# A prospective diagnostic study to measure the accuracy of detection of SARS-CoV-2 Variants Of Concern (VOC) utilising a novel RT-PCR GENotyping algorithm in an In silico Evaluation (VOC-GENIE)

**DOI:** 10.1101/2021.05.05.21256396

**Authors:** Daryl Borley, R.A. Trevor, Alex Richter, Stephen Kidd, Nick Cortes, Nathan Moore, Alice Goring, Kate Templeton, Prachi Teltumbde, Seden Grippon, Paul Oladimeji, Aida Sanchez-Bretano, Andrew Dawson, Joanne E Martin

## Abstract

**Background:** SARS-CoV-2 variants of concern (VOCs) have been associated with higher rate of transmission, and evasion of immunisation and antibody therapeutics. Variant sequencing is widely utilized in the UK. However, only 0.5% (~650k) of the 133 million cumulative positive cases worldwide were sequenced (in GISAID) on 08 April 2021 with 97% from Europe and North America and only ~0.25% (~320k) were variant sequences. This may be due to the lack of availability, high cost, infrastructure and expert staff required for sequencing.

Public health decisions based on a non-randomised sample of 0.5% of the population may be insufficiently powered, and subject to sampling bias and systematic error. In addition, sequencing is rarely available *in situ* in a clinically relevant timeframe and thus, is not currently compatible with diagnosis and treatment patient care pathways. Therefore, we investigated an alternative approach using polymerase chain reaction (PCR) genotyping to detect the key single nucleotide polymorphisms (SNPs) associated with increased transmission and immune evasion in SARS-CoV-2 variants.

**Methods:** We investigated the utility of SARS-CoV-2 SNP detection with a panel of PCR-genotyping assays in a large data set of 640,482 SARS-CoV-2 high quality, full length sequences using a prospective *in silico* trial design and explored the potential impact of rapid *in situ* variant testing on the COVID-19 diagnosis and treatment patient pathway.

**Results:** Five SNPs were selected by screening the published literature for a reported association with increased transmission and / or immune evasion. 344881 sequences contained one or more of the five SNPs. This algorithm of SNPs was found to be able to identify the four variants of concern (VOCs) and sequences containing the E484K and L452R escape mutations.

**Interpretation:** The *in silico* analysis suggest that the key mutations and variants of SARS-CoV-2 may be reliably detected using a focused algorithm of biologically relevant SNPs. This highlights the potential for rapid *in situ* PCR genotyping to compliment or replace sequencing or to be utilized instead of sequences in settings where sequencing is not feasible, accessible or affordable. Rapid detection of variants with *in situ* PCR genotyping may facilitate a more effective COVID-19 diagnosis and treatment patient pathway.

**Funding:** The study was funded by Primer Design (UK), with kind contributions from all academic partners.

## Introduction

In December 2020, the first SARS-CoV-2 variant of concern (VOC) 20I/501Y.V1 (B.1.1.7) was identified in Kent, UK and 20H/501Y.V2 (B.1.3.5.1) was identified in South Africa. Subsequently, VOCs were identified in Brazil and Bristol and more than 20 other significant variants have been identified globally. Variants of concern are associated with higher rate of transmission, mortality and morbidity and/or the potential to evade immunisation and/or antibody therapeutics.^1^B.1.1.7 has been associated with an increased transmission and mortality risk (1.64, 95% CI 1.32 - 2.04)^2^ and vaccines are reported to offer diminished efficacy against variants with the E484K escape mutation.^3^ VOCs with the E484K spike protein mutation include B.1.3.5.1 B.1.351 (South Africa) and VOC-202102/02 (B.1.1.7 with E484K) and P.1 (Brazil) E484K has been described as an escape mutation due to an association with resistance to convalescent sera, antibody therapies and increased re-infection rates.^4^ The E484K and L452R mutations have also been associated with diminished vaccine efficacy, for example the Oxford/AstraZeneca vaccine was reported to be only 21.9% effective against the South African variant^5^ in *in vitro* studies and the Pfizer–BioNTech COVID-19 vaccine elicits antibodies that only partially recognise this variant.^6^

The hospital acquired SARS-CoV-2 infection rate was estimated to be 12.5% in April 2020^7^. Variants with escape mutations are associated with infections with higher mortality rates and the previously used convalescent serum and monoclonal antibodies may not be as effective.^8^ This may lead to an increase in hospital admission and emergency room attendance rates, resulting in a need for rapid *in situ* variant testing to reduce the risk of nosocomial infection with variants. The UK approach to variant testing is a leading continuous nationwide surveillance programme, using genome sequencing^9,10^. Currently, patients’ positive reverse transcription polymerase chain reaction (RT-PCR) samples are reported and then sent to large central sequencing facilities with a turn-around-time of 1-2 weeks. While this timeframe may be sufficient for epidemiological surveillance, this approach is neither patient centric nor able to deliver results in a clinically relevant timeframe. Sequencing is currently not suitable for in situ variant detection due to its inherent lack of speed, on-site availability, and costs. In addition, few nations are meeting the minimum requirements set out by different countries to provide sufficient sample numbers for adequate sequencing based on variant surveillance and there are only ~685k high quality sequences in GISAID^11^, representing just 0.51% of the 133.1 million cumulative positive cases worldwide (as on 08 April 2021) with approximately 97% of those sequences coming from Europe and North America.

An alternative approach utilises RT-PCR genotyping to identify SNPs and was found to provide rapid, cost-effective, and reliable variant monitoring^9^ in a pilot trial. The accuracy of variant PCR-genotyping has not yet been reported with a large data set. Thus, in this study we investigated a large data set of ~640,000 SARS-CoV-2 sequences with PCR-genotyping using a prospective *in silico* trial design, and explored the potential impact on the patient pathways.

## Methods

Five SARS-CoV-2 SNPs were prospectively selected from a review of the published literature for an association with (i) increased SARS-CoV-2 transmission and/or (ii) diminished efficacy of monoclonal antibody therapy, convalescent plasma therapy, vaccine derived immunity, or naturally acquired immunity.10 The study team members that prospectively selected the SNPs were different from those that performed the *in silico* analysis or the investigation of the SNP dataset.

The publicly available Spike Protein Sequence alignment was downloaded from GISAID^11^ on the 19^th^ of March 2021. Subsequently, a further alignment using a multiple sequence alignment program (MAFFT) with a NJ / UPGMA phylogeny was performed. The sequences with more than 1% ambiguous bases were removed from the original dataset of 781,815 sequences. Each protein sequence was then interrogated in excel using the function search string =IF(RIGHT(LEFT(amino acid position, is residue X), 1)=Residue of interest, 1, 0) to determine the number of sequences that contained each SNP of interest. The =IF(AND(Residue X, Residue Y…),1,0) functions were then used to sequentially determine the sequences containing the SNPs, following the algorithm shown in Figure 1 to exclude the Wild type Sequences at each level of the analysis.

**Figure 1.**
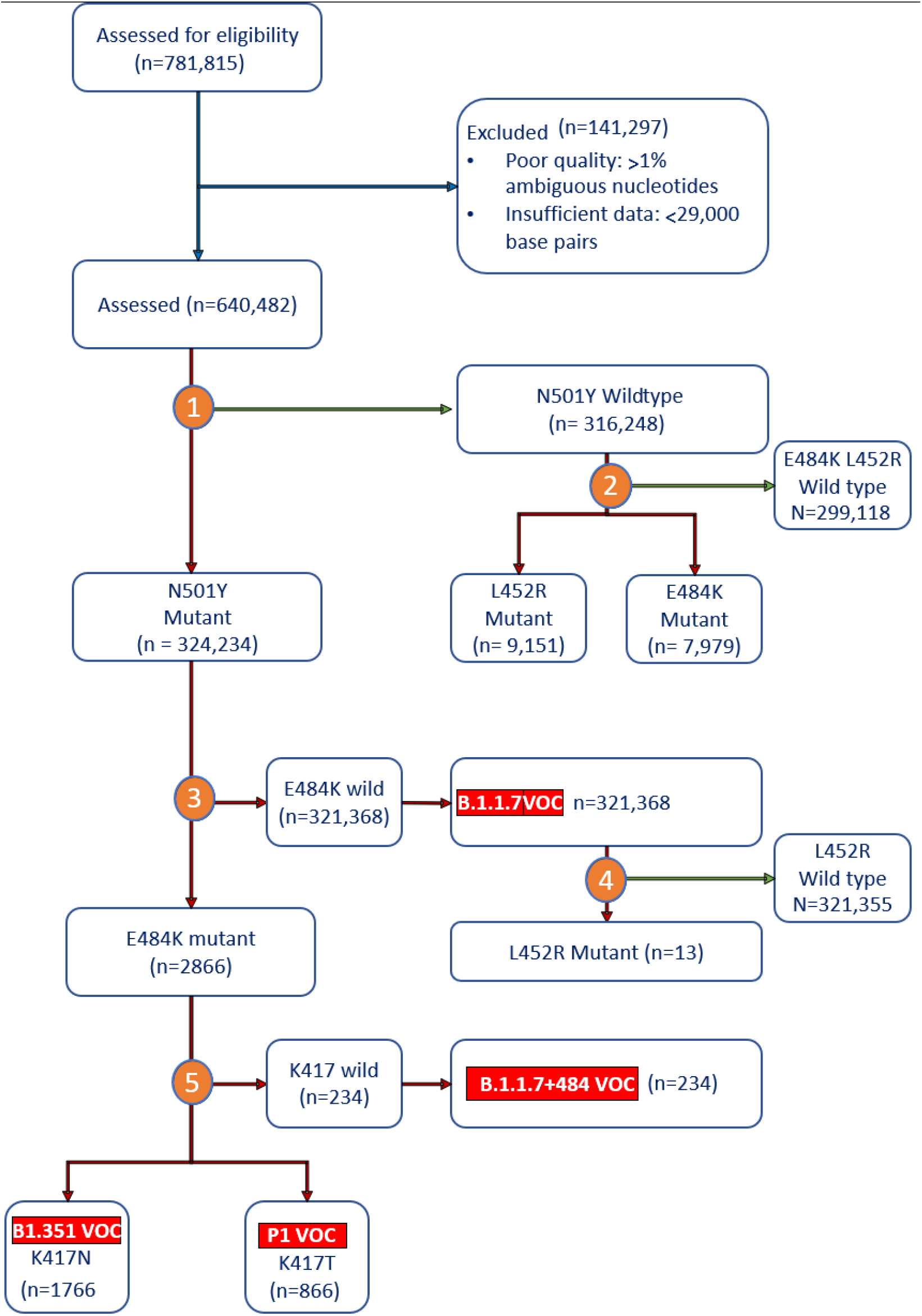
Diagram of the number of GISAID sequences relating to mutations of interest at each step of the algorithm to distinguish VoCs.

**Table 1:** List of the five mutations of interest and their corresponding effect on transmission and immune evasion.

| **SARS-CoV-2 Mutations** | **Increased Transmissibility** | **Evade Antibodies** | **Comments** |
| --- | --- | --- | --- |
| N501Y | Yes <sup>(12-14, 24)</sup> | Yes <sup>(15,16)</sup> | Found in VOCs: B.1.1.7, B.1.351, P.1. Also present in the VUIs P.3, B.1.525, and B.1.526<br>Escapes from cluster 1 antibodies, COVA1-18, COVA2-15, S309 and to a lesser extent, B38 <sup>(15)</sup>. |
| E484K | Yes <sup>(26)</sup> | Yes <sup>(16-18)</sup> | Found in VOCs: B.1.351, P1, VOC-202102/02, and VOI P.2 and P.3<br>Resistant to cluster 2 antibodies <sup>(16,17)</sup><br>Reduces neutralizing activity of human convalescent (2.5- 4.2-fold) and post-vaccination sera <sup>(14,18)</sup><br>VOC B.1.351, P.1 and P.2, which include this mutation, have shown resistance to vaccines <sup>(19)</sup> |
| K417T | No <sup>(26)</sup> | Yes <sup>(16,20,21,25,26)</sup> | Found in VOC P.1<br>Eliminates a salt-bridge between K417 and D30 in ACE2. This salt-bridge is a hotspot of antibody recognition. Its elimination suggests an adaptative role in antibody escape <sup>(20)</sup><br>Escapes from cluster 1 antibodies <sup>(21)</sup> |
| K417N | Moderate <sup>(22)</sup> | Yes <sup>(16, 20-22)</sup> | Found in VOC B.1.351<br>Eliminates a salt-bridge between K417 and D30 in ACE2. This salt-bridge is a hotspot of antibody recognition. Its elimination suggests an adaptative role in antibody escape <sup>(20)</sup><br>The change from K-N creates a new glycosylation site in the protein, considered a classic mechanism to escape antibody binding <sup>(22)</sup>, while glycans have been described as a tool for SARS-CoV-2 to increase infectivity <sup>(23)</sup><br>Escape from cluster 1 antibodies <sup>(21)</sup> |
| L452R | Yes <sup>(25)</sup> | Yes <sup>(25)</sup> | The ability to evade antibodies in the variants B.1.427 and B.1.429 (considered VOC by CDC), has been directly linked, in part, to the presence of L452R <sup>(25)</sup> |

## Results

The five SARS-CoV-2 single nucleotide polymorphisms (SNPs) were selected for their reported association with (i) an increase in SARS-CoV-2 transmission and / or (ii) diminished efficacy of monoclonal antibody therapy, convalescent plasma therapy, vaccine derived immunity, or naturally acquired immunity.^10^

The sequences with more than 1% ambiguous bases were removed from the original dataset of 781,815 sequences. The remaining 640,482 sequences were investigated using the VOC Genie algorithm described in the Figure 1 below: At step (1) 640,482 sequences were assessed for the presence of the N501Y mutation associated with increased transmission. 324,234 sequences contained the SNP while 316,248 were wild type and assessed at step (2) for the E484K and L452R escape mutations. Surprisingly, 7,979 sequences contained the E484K mutation but did not contain the N501Y nor B.1.1.7 identifiers. At step (3), only 2,866 sequences contained the E484K escape mutation while B.1.1.7 was detected in over 321k sequences. The L452R SNP was detected in 13 sequences at (4).

At the (5), B.1.1.7 with E484K (Bristol), South African and Brazil (P1) variants can be distinguished using the K417T/N mutation in combination with the E484K, N501Y and B.1.1.7 SNPs. 234 Bristol VOC sequences were identified, 1,766 South African and 866 Brazilian. The panel of five SNPs detected all the VOCs and mutations of biological significance. The potential impact of rapid PCR identification of variants on patient pathways demonstrated in Figure 2.

**Figure 2:**
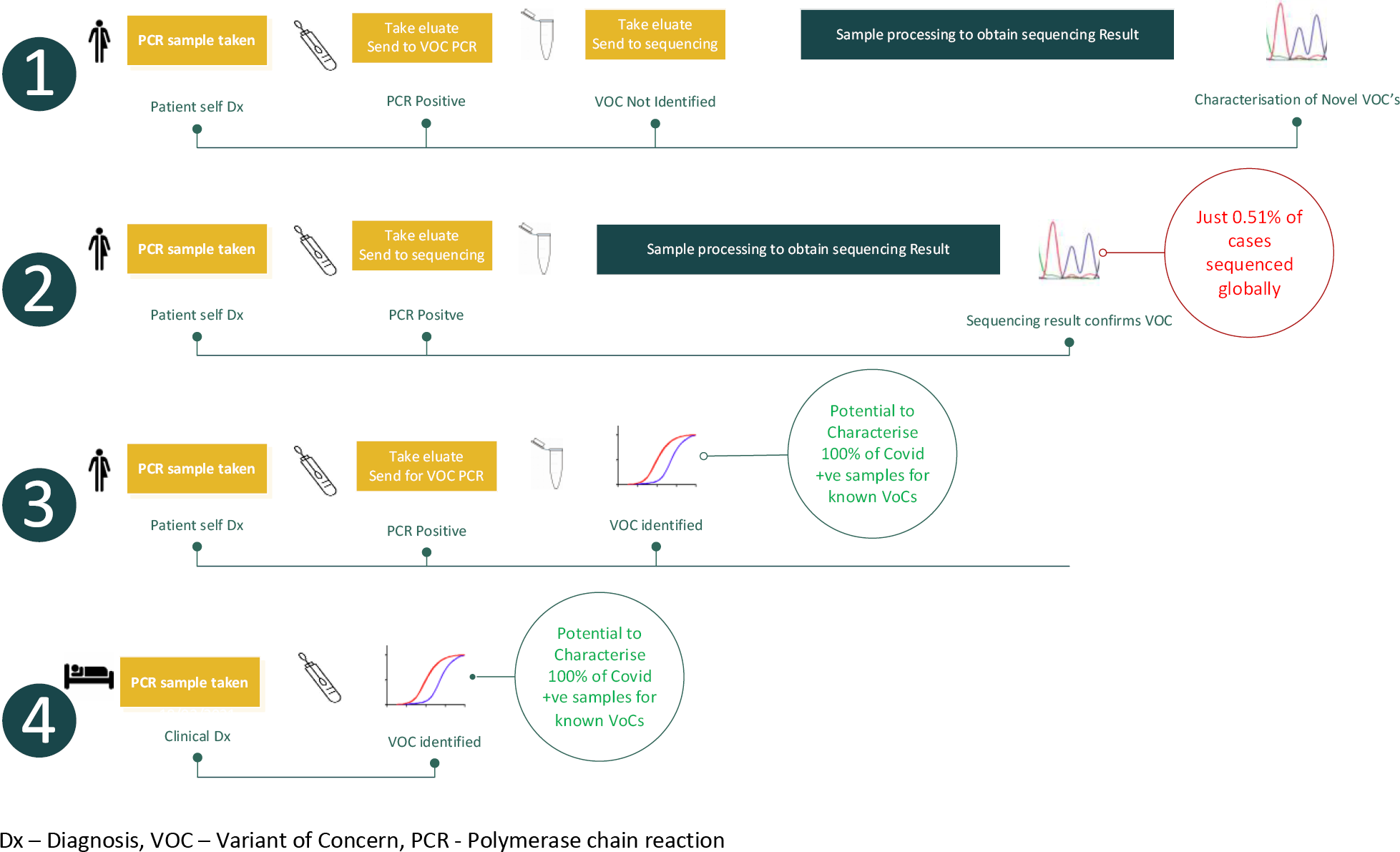
Pathways with simulated timelines for identification of variants used globally.

## Discussion

The SARS-CoV-2 sequences in GISAID represent only 0.5% of the 133 million cumulative cases worldwide, with approximately 97% of those sequences coming from Europe and North America. This is a very small non-randomised sample, which may be insufficiently powered, and subject to sampling bias and systematic error and thus our current understanding of the global epidemiology of SARS-CoV-2 may be extremely limited.

The algorithm of five SNPs identified all four of the variants of concern and mutations of biological significance and two previously unreported SNP combinations (a) N501Y + L452R and (b) E484K in the absence of the N501Y. (Figure 1)

The other potential use of SNP based variant testing is in those cases where high quality nucleic acid is not available for sequencing. 141,297 out of 780,815 cases in GISAID were poor quality or incomplete sequences, for which PCR SNP testing may be a more robust technique. Further, patient samples with VOC genotype also have a higher viral load^27^, meaning these samples are more likely to work for sequencing biasing the results of epidemiological mapping. Accurate genomic sequencing requires a high quality, full length RNA from a sample with a high viral load (~CT<30), whereas due the amplification step, PCR is able to detect VOCs in samples with a low viral load.

From these initial results, PCR genotyping may offer an alternative approach which is cheaper, more accessible and faster alternative to sequencing^9^ and could therefore, be deployed and utilised more broadly to build a more representative data set for public health decision making. Our analysis demonstrates the potential of PCR SNP genotyping to provide rapid *in situ* variant detection with a widely accessible and clinically relevant approach.

PCR genotyping is limited to SNPs that have already been identified by sequencing, and so sequencing is still required for *de novo* detection. However, the public health and clinical interventions are not based on *de novo* detection but on the evidence that a known variant has a deleterious adaption. Thus, PCR genotyping maybe better suited for clinical and population variant detection, allowing the scarce sequencing resources to be better utilised for *de novo* detection of biologically relevant SNPS. The potential changes to the COVID-19 patient pathway from including PCR genotyping are explored in Figure 2 below:

1. PCR and then VOC PCR where novel variants could be identified for sequencing.
2. Current Pathway: PCR and then sequencing to identify VOCs
3. Positive PCR reflexed on to VoC PCR for variant identification
4. Clinically diagnosed hospital patients tested directly for variants

Usually positive COVID-19 tests are sent to a network of specialised centres for sequencing and to determine the presence of SNPs and VOCs. This is the traditional pathway 2 in figure 2 above and is associated with delay in availability of results, increased costs as well as the need for more samples from individual patients. Alternatively in pathway 3, the PCR VOC pathway, a single positive sample is used to detect VOCs and the key SNPs in approximately two hours. Pathway 1 is the most efficient and effective in settings where sequencing is available – samples follow the PCR VOC pathway and only samples that don’t match known VOCs and SNP combinations i.e., potential *de novo* variants are sent for sequencing.

In the hospital setting, pathway 4, the PCR detection of VOCs can be used in a near patient setting to facilitate testing and treatment decisions in a clinically relevant timeframe and also to test all staff and patients attending or being admitted to hospitals to reduce the risk of nosocomial transmission. This pathway is also applicable to the office, transport, educational and large event settings, which have acted as super spreader events for SARS-CoV-2 and perhaps exemplifies the potential for a simple and reliable rapid PCR technology might be deployed to underpin a ‘return to normal’ economic recovery.

## Data Availability

The data can be made available on request

## Contributorship statement

Contributors: All authors (DB, RAT, PT, SG, PO, ASB, AR, SK, NM, NC, AG, AP, HM, AD, JM) contributed to designing the work, analysing the data, and drafting and revising the manuscript.

## Acknowledgement

Tom Jefferson contributed to data acquisition.

## Declaration of interests (please fill in against name)

Stephen Kidd, Nick Cortes, Nathan Moore, Kate Templeton, Alex Richter and Alice Goring have no conflicting interests.

R.A Trevor, Daryl Borley, Paul Oladimeji, Prachi Teltumbde, Seden Grippon, Andrew Dawson and Aida Sanchez-Bretano are employees of Novacyt group, which is a medical diagnostics company operating in the COVID-19 variant testing field. R.A Trevor has no additional direct conflicts but is a shareholder in a number of un-related private and public companies that do not operate in the COVID-19 or diagnostics field.

Joanne Martin has no direct conflicts of interest. She is a principal investigator of a care home trial using Novacyt rapid testing and National Specialty Advisor for Pathology for NHS England and Improvement. She is a director and shareholder of Biomoti a drug delivery company and has a shareholding in Glyconics, a diagnostics company.

